# Responsible modelling: Unit testing for infectious disease epidemiology

**DOI:** 10.1101/2020.08.14.20175216

**Authors:** Tim CD Lucas, Timothy M Pollington, Emma L Davis, T Déirdre Hollingsworth

**Author notes:** Email address (Tim CD Lucas).

## Abstract

Infectious disease epidemiology is increasingly reliant on large-scale computation and inference. Models have guided health policy for epidemics including COVID-19 and Ebola and endemic diseases such as malaria and tuberculosis. Yet a single coding bug may bias results, leading to incorrect conclusions and wrong actions that could cause avoidable harm. We are ethically obliged to ensure our code is as free of error as possible. Unit testing is a coding method to avoid such bugs, but unit testing is rarely used in epidemiology. We demonstrate through simple examples how unit testing can handle the particular quirks of infectious disease models.

## 1. Introduction

Modelling is an important tool for understanding fundamental biological processes in infectious disease dynamics, evaluating potential intervention efficacy and forecasting disease burden. At the time of writing, infectious disease modellers are playing a central role in the interpretation of available data on the COVID-19 pandemic to inform policy design and evaluation [1–3]. Similarly, policy on endemic infectious diseases, such as duration and frequency of control programmes and spatial prioritisation, is also directed by models [4]. Such research builds on a long history of modelling for policy [5] and a general understanding of the dynamics of infectious disease systems.

Given the importance of modelling results, it is vital that the code they rely on is both coded correctly and trusted. Bugs can be caused by typos, code behaving in unexpected ways, or logical flaws in the construction of the code. Outside of epidemiology, bugs have been found in code that had been used by many researchers [6] and may lead to retractions [7]. Bugs have also been found in highly influential work; a paper that informed austerity policies globally was found to have a fatal computational mistake [8]. In engineering bugs caused the Mars Climate orbiter and the Mariner 1 spacecraft to become lost or destroyed [9,10]. We do not know of high profile cases of infectious disease models being found to have bugs once published, but as code is not always shared and little post-publication testing of code occurs, this likely represents a failure of detection. The issue of trust was highlighted recently when Neil Ferguson, one of the leading modellers informing UK COVID-19 government policy, tweeted:

> “I’m conscious that lots of people would like to see and run the pandemic simulation code we are using to model control measures against COVID-19. To explain the background – I wrote the code (thousands of lines of undocumented C) 13+ years ago to model flu pandemics…” [11].

The code that was released did not include any tests [12] but subsequent work has added documentation, while independent code reviews have supported the results of the study [13,14]. The tweet and lack of tests garnered considerable backlash (some of which may have been politically motivated [15]), with observers from the software industry noting that code should be both documented and tested to ensure its correctness [14]. It is understandable that during the fast-moving, early stages of a pandemic, other priorities were put above testing and documenting the code. However, this simply reinforces that time should be taken to test and document code when it is first written so that it is already well tested for when it is needed under short deadlines. It is also important to note that a lack of tests is not unusual in our field, or for some of the authors of this article. To guard against error, policy-makers now standardly request analyses from multiple modelling groups (as is the case in the UK for COVID-19 [16]) as a means to provide scientific robustness and reliability [17], yet this is not enough if the models themselves lack internal validity.

Infectious disease modellers are rarely trained as professional programmers [14] and recently some observers have made the case that this has been due to a lack of funding [18]. It is also notable that there are few texts available which demonstrate the use of unit testing to check infectious disease models. Infectious disease modellers have sought to address these issues (as in the case above) by having multiple, independently developed models to inform policy, to address structural and parameter uncertainty [17,19].

Whilst there many drivers and attempts to address this problem with code robusteness, today’s models are increasingly moving from mean-field ordinary differential equation approximations to individual-based models with complex, data-driven contact processes [20,21]. These increases in model complexity are accompanied with growing codebases. As the mathematical methods depend increasingly on numerical solutions rather than analytical pen-and-paper methods, it becomes more difficult to tell if a bug is present based on model outputs alone.

Traditionally, the workflow is to write the complete code, run the model and examine plots of the output. This crude *ad hoc* testing approach can miss many bugs. Furthermore, this workflow is biased as models that show expected behaviour are assumed bug-free, whereas the opposite gets more scrutiny.

*Unit testing* is a formally-defined, principled framework that compares comprehensive output scenarios from code to what the programmer expected to happen ([22] Chapter 7, [23], [24]). Ready-to-run frameworks for unit testing are available in *R* [25], *Julia* [26] and *python* [27] and are standard practice in the software industry. These testing concepts also apply to many other scientific fields but here we focus on infectious diseases. Infectious disease modelling presents specific challenges, such as stochastic outputs, which are difficult to test and not covered in general unit testing literature.

In this primer we introduce unit testing with a demonstration of an infectious disease model. We also outline the available testing frameworks in various languages commonly used by modellers.

## 2. Unit testing foundations

At the heart of every *unit test* is a function output, its known or expected value and a process to compare the two. For the square root function (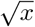 or `sqrt(x)` in *R*), we could write a test that runs the function for the number 4, i.e. `sqrt(x = 4)`, and compares it to the correct answer i.e. 2. However, often our function arguments will cover an infinite range of possibilities and we cannot exhaustively check them all. Instead we devise tests that cover standard usage as well as *corner case* scenarios: what do we want our function to do if given a negative number e.g. `sqrt(−1)`, or a vector argument containing strings or missing values e.g. `sqrt(c(4,”melon”,NA))`?

In R, the `testthat` package [28], provides a simple interface for testing. While a variety of test functions can make different comparisons, the two main ones are `expect_true()` and `expect_equal()`. `expect_true()` takes one argument: an expression that should evaluate to `TRUE`. For our square root example above, we would write `expect_true(sqrt(4) = = 2)`. `expect_equal()` takes two arguments, an expression and the expected output; so we would write `expect_equal(sqrt(4), 2)`.

There are a number of ways to incorporate unit testing into your programming workflow.

1. Each time you write code for a new, discrete chunk of functionality, you should write tests that confirm it does what you expect. These tests should be kept in the same directory as the code it is testing.
2. Whenever a bug is found in the code outside of the existing testing framework, a new test should be written to capture it. Then if the bug re-emerges it will hopefully be quickly flagged so that the developor can fix it.
3. All of these tests should be regularly run as you develop new code. If a change causes the older tests to break, this points to the introduction of an error in the new code, or implies that the older code could not generalise to the adapted environment.

## 3. An example multi-pathogen re-infection model

Here we define a toy epidemiological model and then demonstrate how to effectively write unit tests for it in *R* code. Consider a multi-pathogen system, with a population of *N* infected individuals who each get infected by a new pathogen at every time step (Fig. 1). In this toy example, we imagine that individuals are infected with exactly one pathogen at a time. Some aspects of this model could reflect the dynamics of a population where specific antibiotics are used regularly i.e. each time step an individual is infected, diagnosed and treated suboptimally, leaving the individual susceptible to infection from any pathogen, including the one they were just treated for. The aim of this model however is not to be realistic but serve as a learning tool with succinct code.

**Figure 1:**
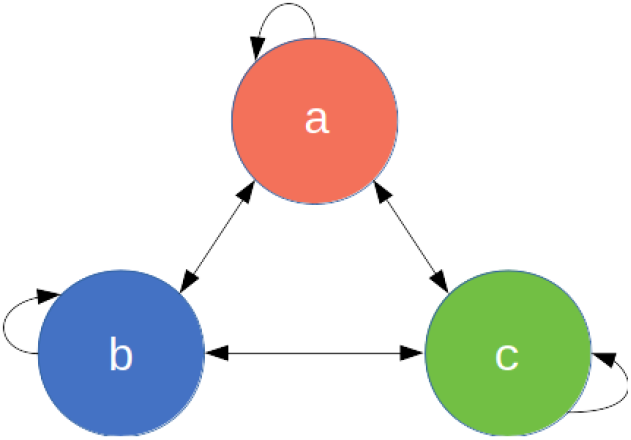
The 3-pathogen system with arrows showing the possible transitions at every time step.

Each individual *i*, at time *t*, is defined by the pathogen they are currently infected with *I_it_* ∈ *{a, b, c}* for a 3-pathogen system. The population is therefore defined by a length *N* state vector **I**_t_ = (*I*_*it*_)_*i* = [*1,N*]_. At each time step, every individual’s infection status is updated as:

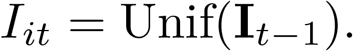

That is, at each iteration, the new infection status of each individual is a Uniform random sample from the set of infection statuses in the previous iteration (including itself *I*_*i,t*−1_). Certainly this toy model is naïve as it is governed by mass-action principles, ignoring contact and spatial dynamics. Nevertheless it will serve its purpose. Code 1 shows our first attempt at implementing this model.

#### Code 1: Base example of the multi-pathogen re-infection model

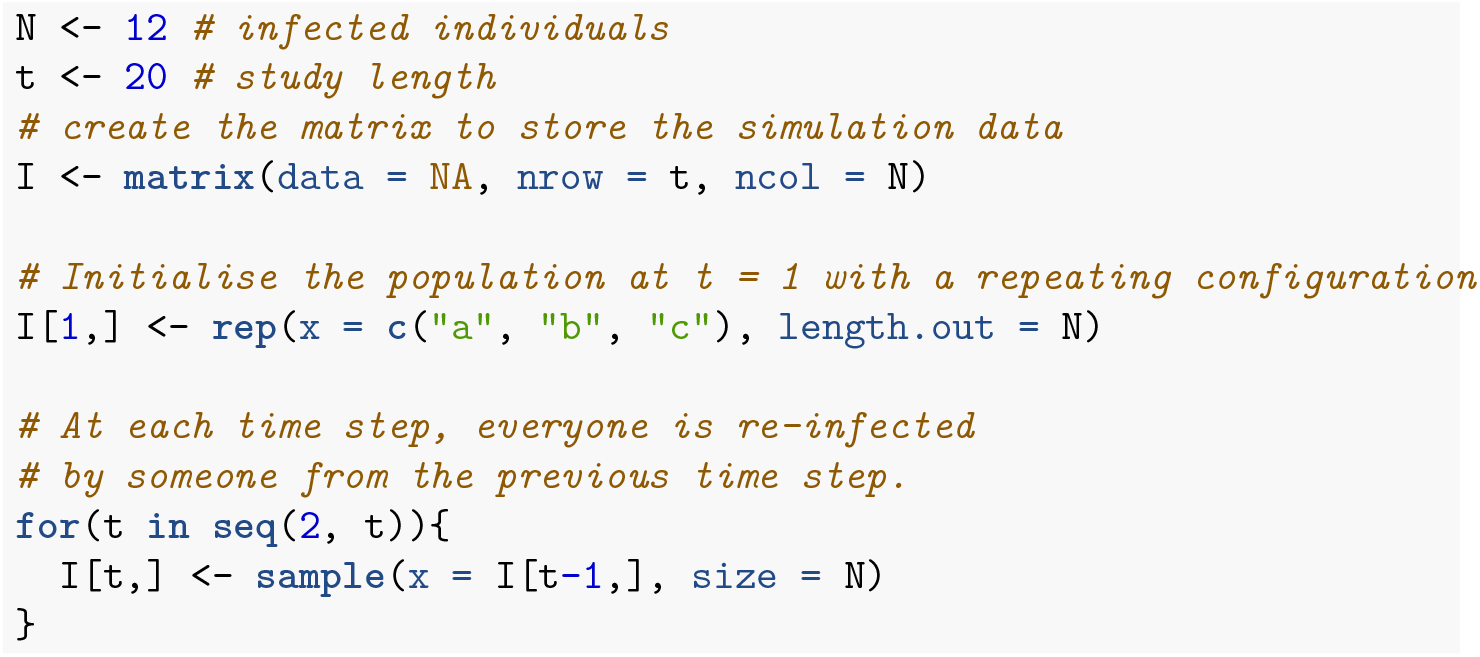

Usually we would make some output plots to explore if our code is performing sensibly. Plotting the time course of which pathogen is infecting one individual shows repeated infection with different pathogens as expected (Fig. 2). However, if we look at the proportion of each pathogen through time (not shown here) we quickly see that the pathogen proportions are identical through time and so there must be a bug that we had not originally noticed. This simple example demonstrates a number of points. Firstly, bugs can be subtle. Secondly, it is not easy to notice an error, even in just 7 lines of code. Thirdly, it is much easier to debug code when you know there is a bug. Fourthly, while plotting simulation runs is an excellent way to check model behaviour, if we had only relied on Fig. 2 we would have missed the bug. Additionally, manually checking plots is a time consuming and non-scalable method because a human has to perform this scan every test run. In summary this *ad hoc* plotting approach reduces the chances that we will catch all bugs.

**Figure 2:**
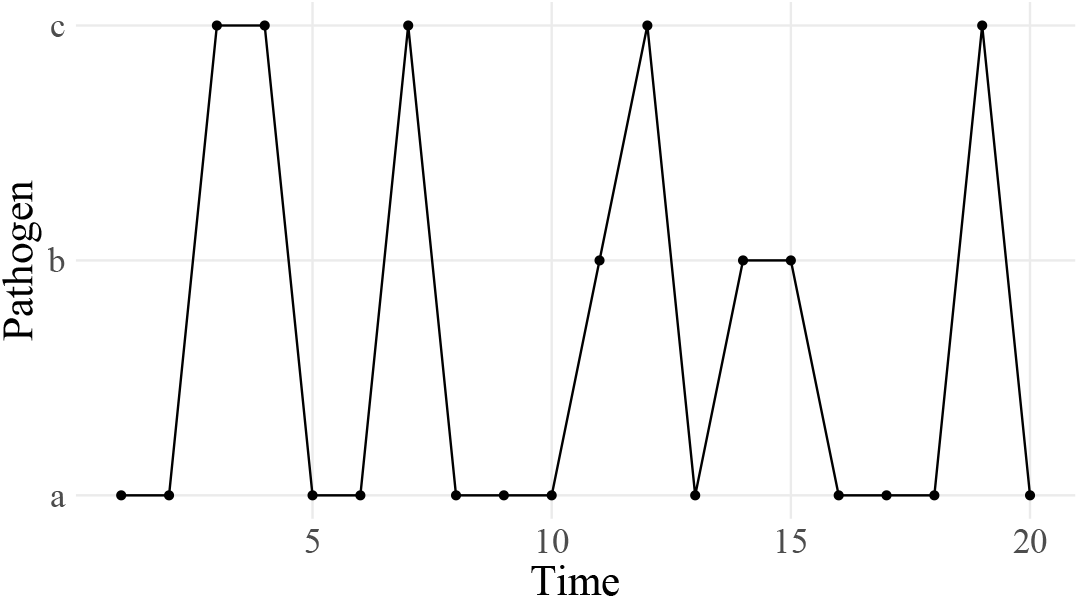
Infection profile for individual 1, who is initially infected with pathogen *a*.

The cause of the bug is that `sample()` defaults to sampling without replacement `sample(…, replace = FALSE)`; this means everyone transmits their infection pathogen on a one-to-one basis rather than one-to-many as required by the model. Setting `replace = TRUE` fixes this (Code 2) and when we plot the proportion of each pathogen (Fig. 3) we see the correct behaviour (a single pathogen drifting to dominance). In the subsequent sections we will develop this base example as we consider different concepts in unit testing, resulting in well-tested code by the end.

#### Code 2: Corrected base example

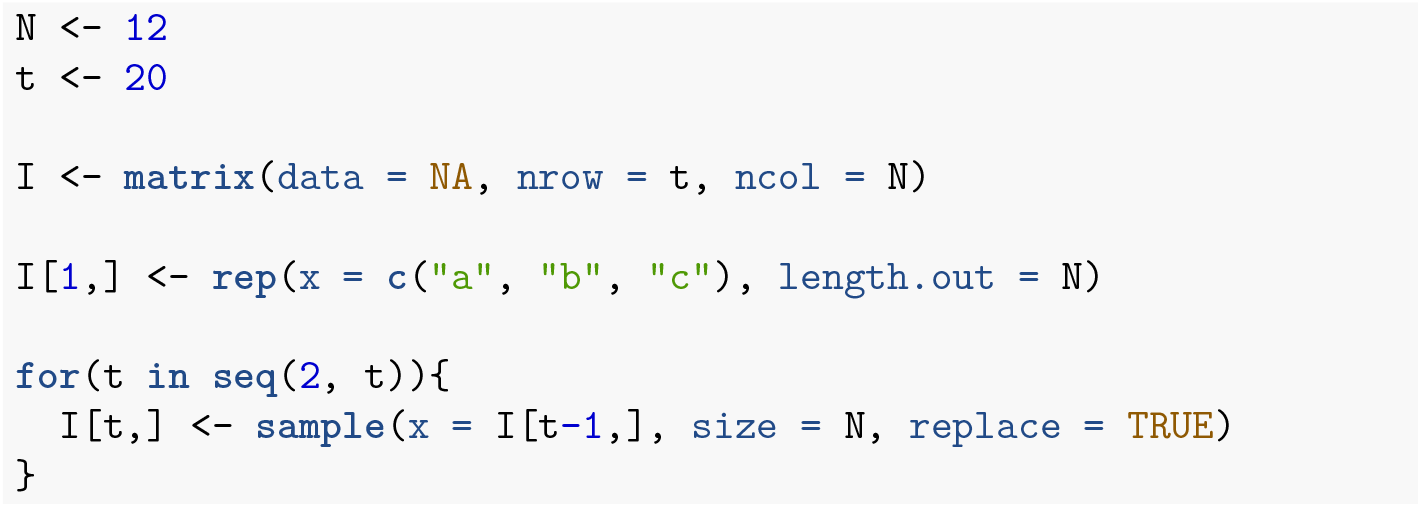

**Figure 3:**
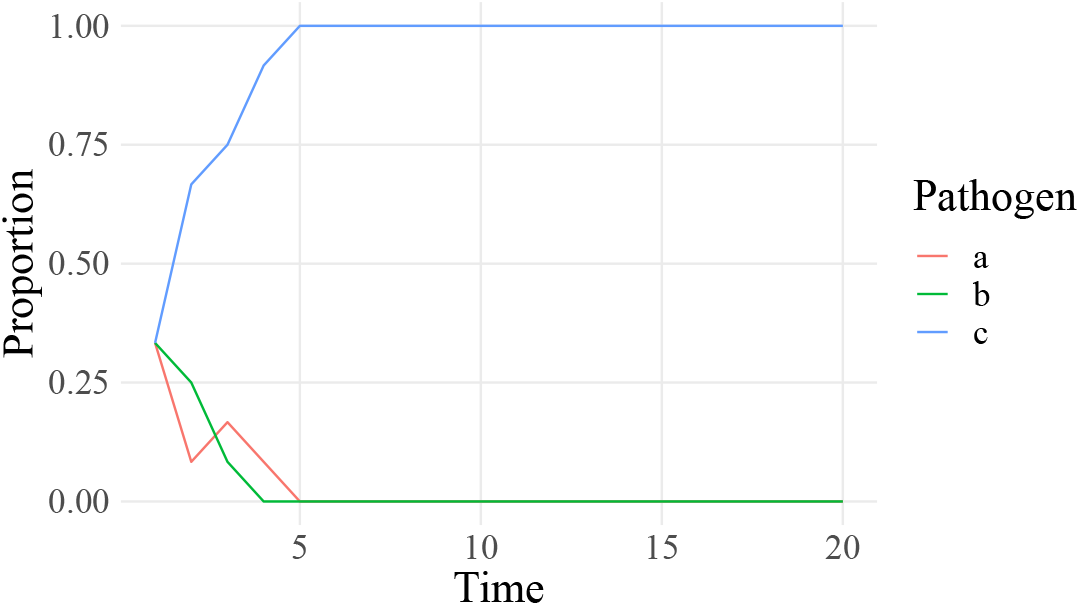
The correct behaviour with the proportion of each pathogen drifting to either dominance or extinction. Each pathogen is a different line.

## 4. Basic unit testing

### Write small functions

To ensure the unit tests are evaluating the exact code as run in the analysis, code should be structured in functions, which can be used to both run unit tests with and to generate results as part of a larger model codebase. Make your functions compact with a single clearly-defined task. We have defined a function, `initialisePop()`, to initialise the population and another, `updatePop()`, to run one iteration of the simulation (Code 1). Organising the codebase into these bite-sized operations makes following the programming flow easier as well as understanding the structure of the code. At this stage we have also enabled the varying of the number of pathogens using the `pathogens` argument in the `initialisePop()` function. The first iteration of the simulation, `I[1,]`, is initialised with a repeating sequence of letters.

#### Code 1: Organising code into small functions

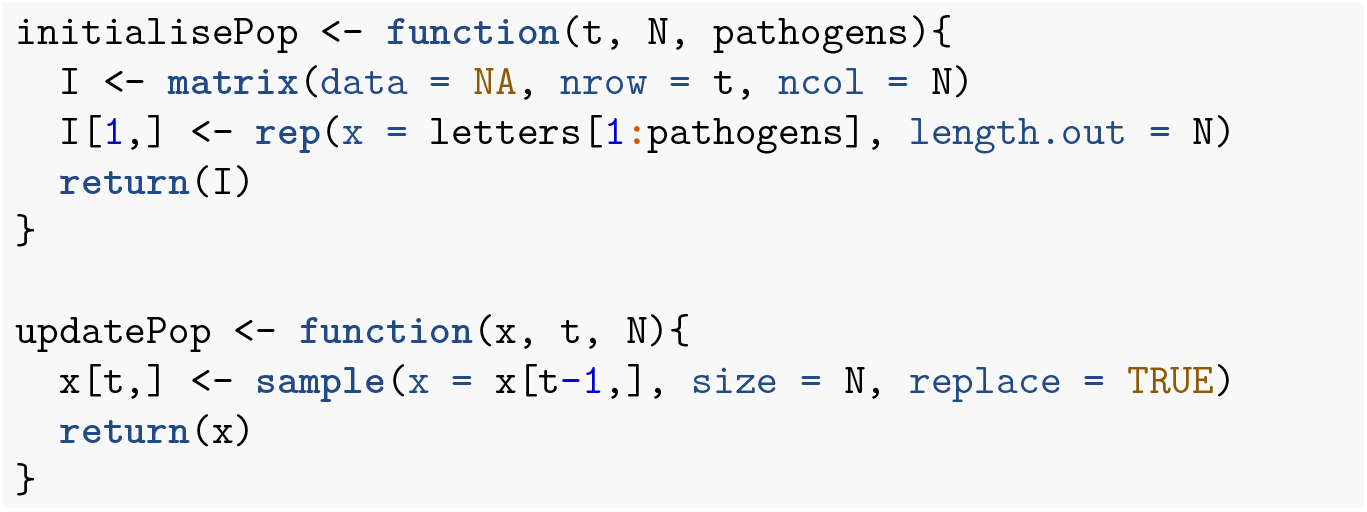

### Test simple cases first

If we start with a small population with few pathogens, we can then easily work out exactly what the initialised population should look like (Code 2). When we initialise a population with three individuals and three pathogens, we will get the sequence “a”, “b”, “c” as seen in the first test. When the number of individuals is greater than the number of pathogens, the sequence will be repeated as seen in the second test. Finally, when the number of individuals is greater than the number of pathogens, but is not a multiple of the number of pathogens, the sequence will have an incomplete repeat at the end as seen in the third test. In this sequence of tests, we have taken our logical understanding of what the function should do, and used it to make predictions of what the results should be. We then test that the result is the same as what we expect.

#### Code 2: Using simple parameter sets we can work out beforehand what results to expect

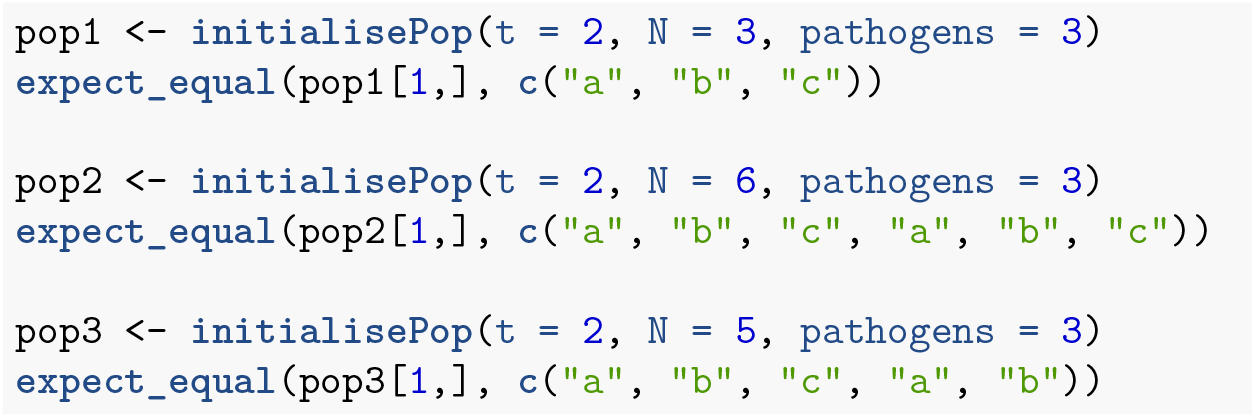

### Test all arguments

`initialisePop()` has three arguments to check. First we initialise the population, and then alter each argument in turn (Code 3). Arguments `t` and `N` directly change the expected dimension of the returned matrix so we check that the output of the function is the expected size. For the `pathogens` argument we test that the number of pathogens is equal to the number requested.

#### Code 3: Test all function arguments in turn

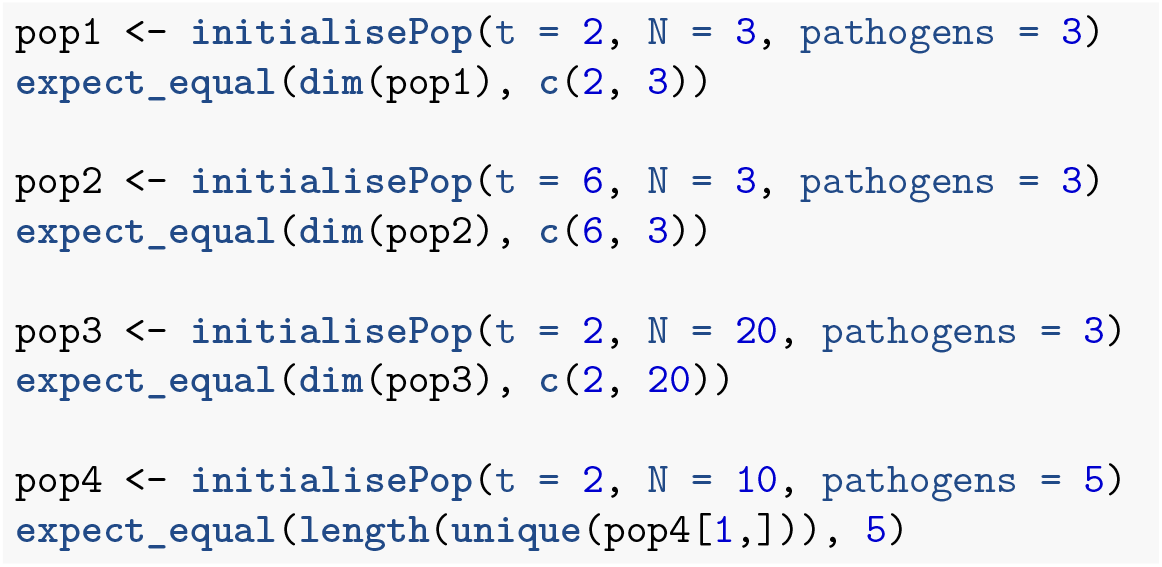

### Does the function logic meet your expectations?

We can also cover cases that expose deviations from the logical structure of the system. After initialising our population, we expect all the rows other than the first to contain `NA`. We also expect each of the pathogens *a, b* and *c* to occur at least once on the first row if `pathogens` = 3 and `N` ≥ 3. Finally, `updatePop()` performs a single simulation time step, so we expect only one additional row to be populated. Instead of testing by their numerical values, we verify logical statements of the results within our macro understanding of the model system (Code 4).

#### Code 4: Test more complex cases using your understanding of the system

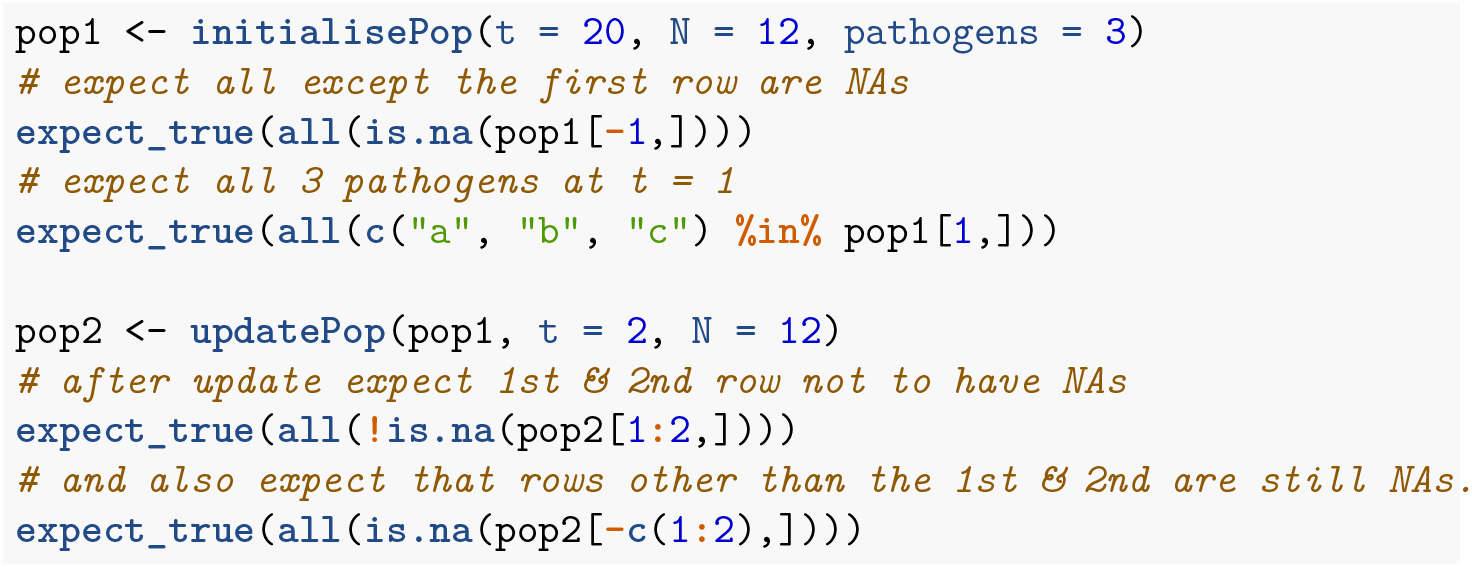

### Combine simple functions and test them at a higher-level

In the end an entire model only runs when its functions work together seamlessly. So we next check their connections; achieved through nesting functions together, or defining them at a higher level and checking the macro aspects of the model. We define a function `fullSim()` that runs both `initialisePop()` and `updatePop()` to yield one complete simulation. We would expect there to be no `NA`s in the output from `fullSim()` and every element to be either *a, b* or *c*.

## 5. Stochastic code

Stochastic simulations are a common feature in infectious disease models. Stochastic events are difficult to test effectively because, by definition, we do not know beforehand what the result will be. We can check very broad-scale properties, like Code 5, where we check the range of pathogen values. However, code could still pass and be wrong (for example the base example (Code 1) would still pass that test). There are however a number of approaches that can help.

#### Code 7: Combine simple functions through nesting to check higher-level functionality

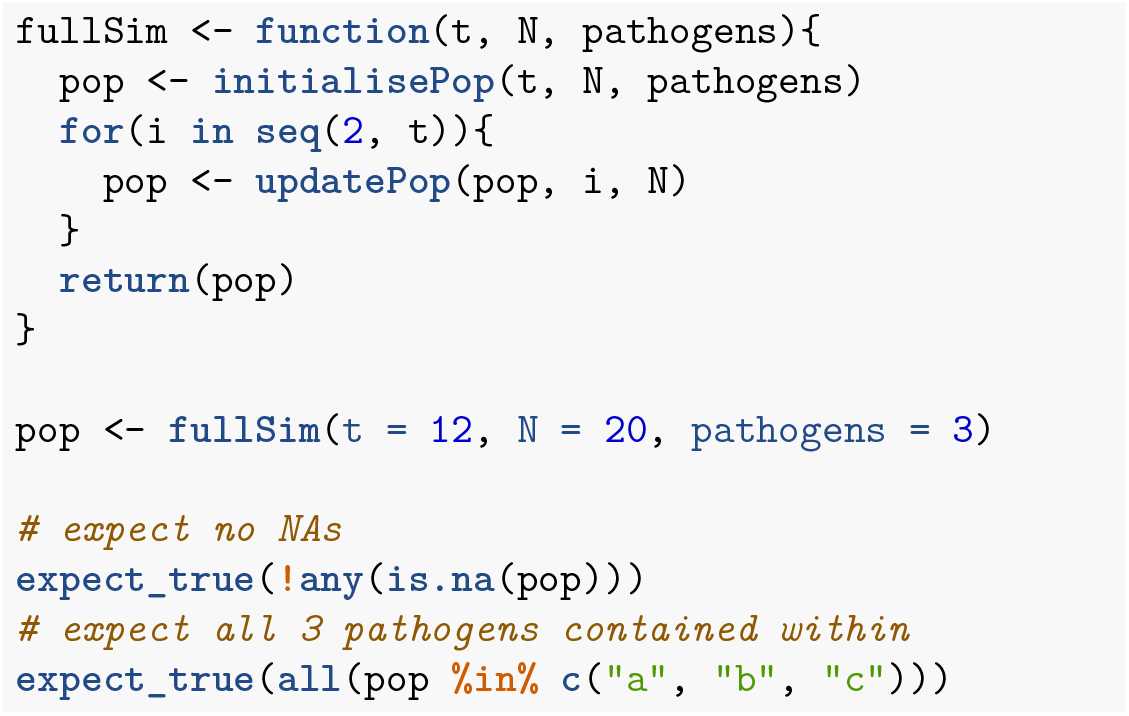

### Split stochastic and deterministic parts

Isolate the stochastic parts of your code. For example, `updatePop()` performs stochastic and deterministic operations in one line (Code 1). Firstly, `updatePop()` stochastically samples who gets infected by whom at iteration `t`. Then it takes those infection events and assigns the new infectious status for each individual. We demonstrate in Code 1 how this could be split. We accept this is a fairly exaggerated example and splitting a single line of code into two functions is rare! The more common scenario is splitting a multi-line function into smaller functions which also brings benefits of code organisation so it does not feel like extra effort.

#### Code 1: Isolation of the determistic and stochastic elements

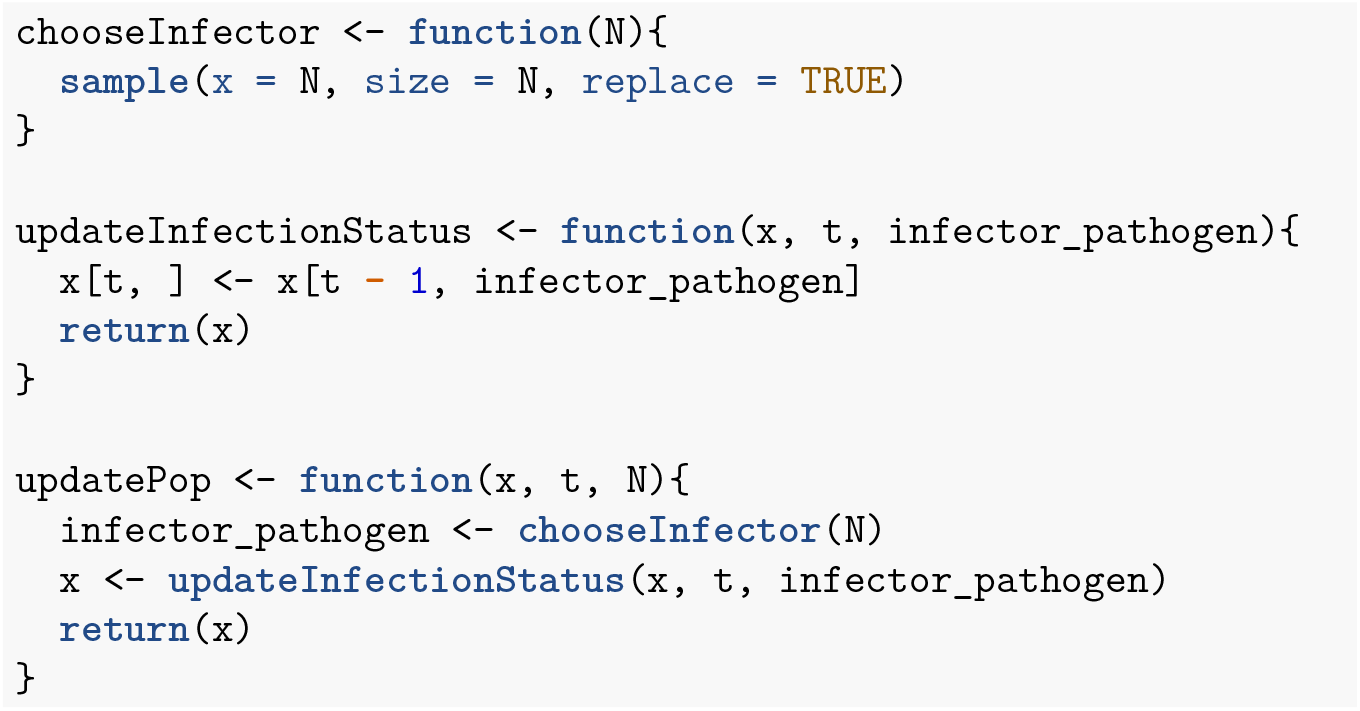

Now, half of `updatePop()` is deterministic so can be checked as previously discussed. We still have `chooseInfector()` that is irreducibly stochastic. We now examine some techniques for directly testing the stochastic parts of a model.

### Pick a smart parameter for a deterministic result

In the same way that we used simple parameters values in Code 2, we can often find simple cases for which our stochastic functions become deterministic. For example, samples from *X* ∼ Bernoulli (p) will always be zeroes for *p* = 0 or ones for *p* = 1. In the case of a single pathogen (Code 2), the model is no longer stochastic. So initialisation with one pathogen means the second time step should equal the first.

#### Code 2: A stochastic function can output deterministically if you can find the right parameter set.

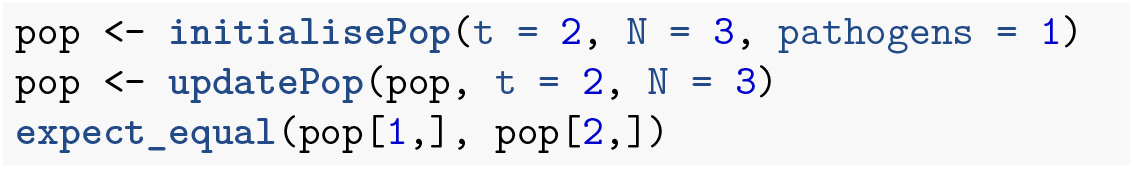

### Test all possible answers (if few)

Working again with a simple parameter set, there are some cases where the code is stochastic, but with a small, finite set of outputs. So we can run the function exhaustively and check it returns all of the possible outputs. For a population of two people, `chooseInfector()` returns a length-2 vector with the possible elements of 1 or 2. There are four possibilities when drawing who is infected by whom. Both individuals can be infected by individual 1, giving the vector {1, 1}. Both individuals can be infected by individual 2, giving {2, 2}. Both individuals can infect themselves, giving {1, 2}. Or finally both individuals can infect each other, giving {2, 1}. In (Code 3), `chooseInfector(N = 2)` returns a length-2 vector with the indices of the infector for each infectee. `paste0()` then turns this length-2 vector into a length-1 string with two characters; we expect this to be one of “11”, “22”, “12” or “21”. `replicate()` runs the expression 300 times, but in your unit test you should choose a value high enough so that you are confident that all of the distinct outcomes will have occurred at least once.

#### Code 3: Testing stochastic output when it only covers a few finite values

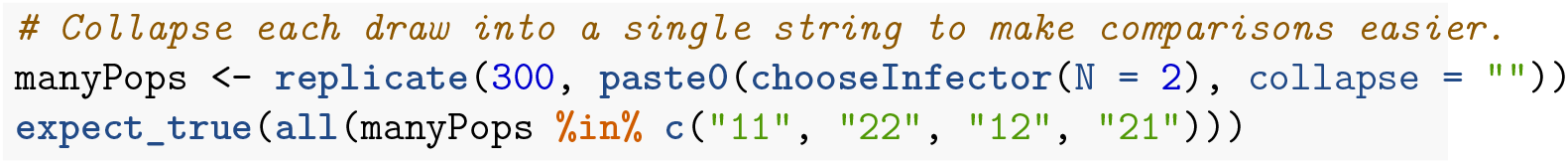

### Use very large samples for the stochastic part

While the previous example worked well for a small set of possible outputs, testing can conversely be made easier by using very large numbers. This typically involves large sample sizes or numbers of stochastic runs. For example, the clearest test to distinguish between our original, buggy code (Code 1) and our correct code (Code 2) is that in the correct code there is the possibility for an individual to infect more than one individual in a single time step. In any given run this is never guaranteed, but the larger the population size the more likely it is to occur. With a population of one thousand, the probability that no individual infects two others is vanishingly rare (Code 4). As this test is now stochastic we should set the seed of the random number generator so that the test is reproducible. Setting the seed with `set.seed` means that each time the code is run, the same pseudo-random numbers will be generated.

#### Code 4: Testing that the code does allow one individual to infect multiple individuals.

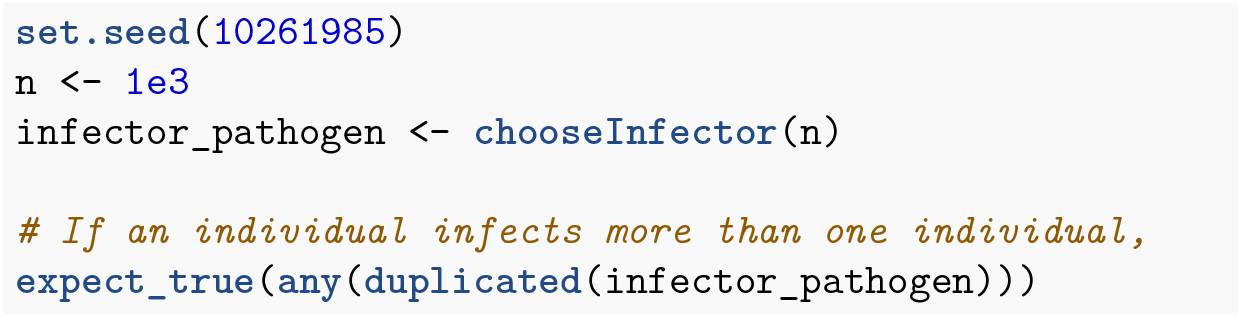

If we have an event that we know should never happen, we can use a large number of simulations to provide stronger evidence that it does not stochastically occur. However, it can be difficult to determine how many times is reasonable to run a simulation, especially if time is short. This strategy works best when we have a specific bug that occurs relatively frequently (perhaps once every ten simulations or so). If the bug occurs every ten simulations and we have not fixed it we would be confident that it will occur at least once if we run the simulation 500 or 1000 times. Conversely, if the bug does not occur even once in 500 or 1000 simulations we can be fairly sure we have fixed it.

Similarly, a bug might cause an event that should be rare to happen very regularly or even every time the code is run. In our original buggy code (Code 1) we found that the proportions remained identical for entire simulations. We would expect this to happen only very rarely. We can run a large number of short simulations to check that this specific bug is not still occurring by confirming that the proportion of each pathogen is not always the same between the first and last time point. As long as we find at least one simulation where the proportions of each pathogen are different between the first and last iteration, we know the bug has been fixed.

#### Code 5: Assessing if a bug fix was a likely success with large code runs, when the bug was appearing relatively frequently

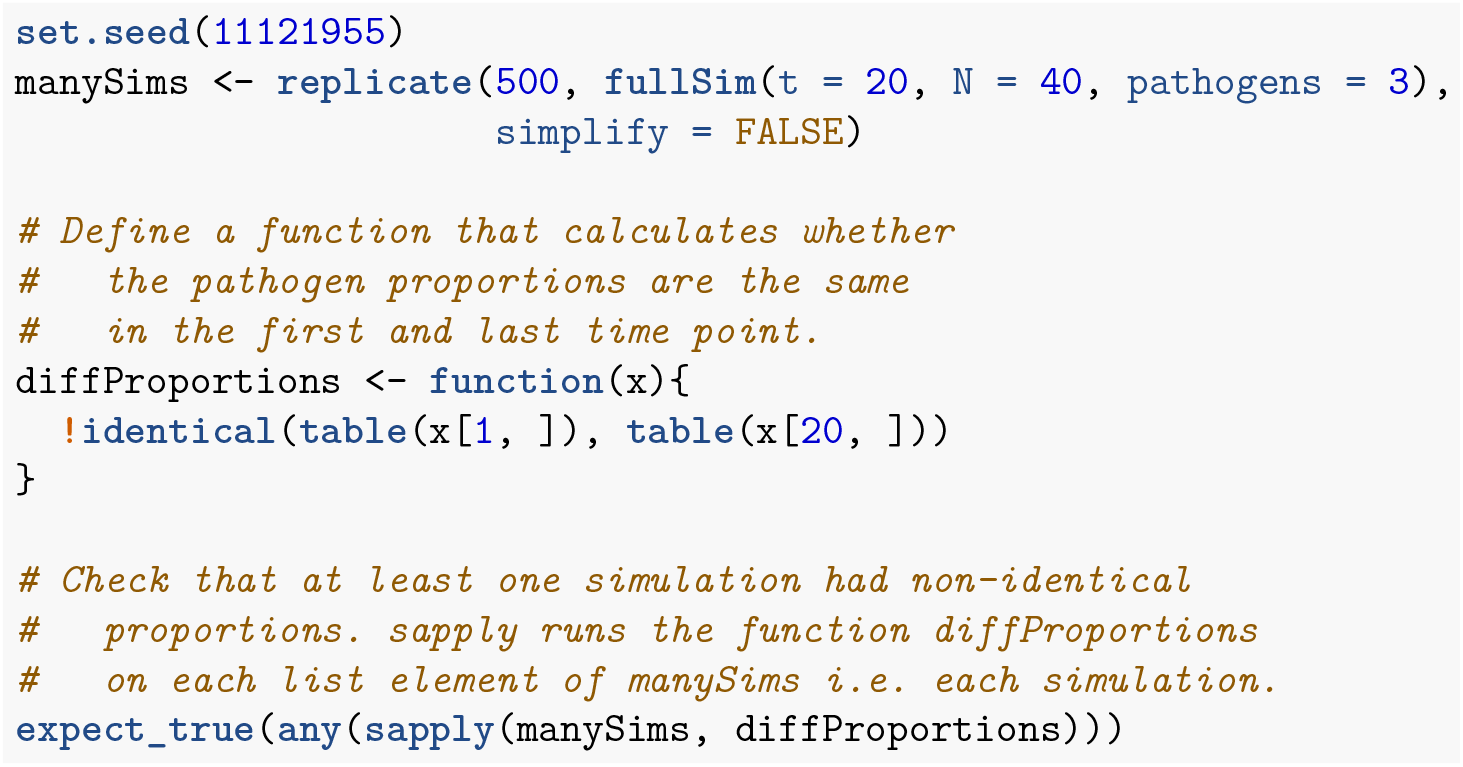

## 6. Further testing

### Test incorrect inputs

As well as testing that functions work when given the correct inputs, we must also test that they behave sensibly when given wrong ones. This typically involves the user inputting argument values that do not make sense. This may be, for example, because the inputted argument values are the wrong class, in the wrong numeric range or have missing data values. Therefore it is useful to test that functions fail gracefully if they are given incorrect inputs. This is especially true for external, exported functions, available to a user on a package’s front-end. However, it is not always obvious what constitutes an ‘incorrect value’ even to the person who wrote the code. In some cases, inputting incorrect argument values may cause the function to fail quickly. In other cases code may run silently giving false results or take a long time to give an error. Both of these cases can be serious or annoying and difficult to debug afterwards.

Often for these cases, the expected behaviour of the function should be to give an error. There is no correct output for an epidemiological model with –1 pathogens. Instead the function should give an informative error message. Often the simplest solution is to include argument checks at the beginning of functions. We then have to write slightly unintuitive tests for an expression where the expected behaviour is an error. If the expression does not throw an error the test should throw an error (as this is not the expected behaviour). Conversely, if the expression does throw an error the test should pass and not throw an error. We can use the `expect_error()` function for this task. This function takes an expression as its first argument and reports an error if the given expression does not throw an error as expected.

We can first check that the code sensibly handles the user inputting a string instead of an integer for the number of pathogens. Because this expression throws an error, `expect_error()` does not throw an error and the test passes.

#### Code 1: Testing incorrect pathogen inputs

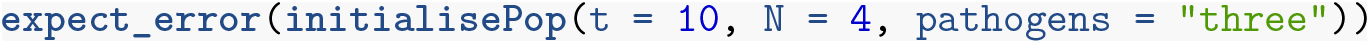

Now we contrast what happens if the user inputs a vector of pathogens to the `initialisePop()` function. Here we are imagining that the users intent wass to run a simulation with three pathogens: 1, 2 and 3.

#### Code 2: A failing test for incorrect pathogen inputs

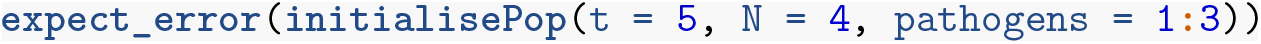

This test fails because the function does not throw an error. Instead the code takes the first element of `pathogens` and ignores the rest. Therefore, a population is created with one pathogen, not three, which is almost certainly not what the user wanted. Here, the safest fix is to add an explicit argument check at the top of the function as implemented below. The same test now passes because `initialisePop()` throws an error when a vector is supplied to the `pathogens` argument.

#### Code 3: New definition of the initialisePop() function and a passing test for incorrect pathogen inputs

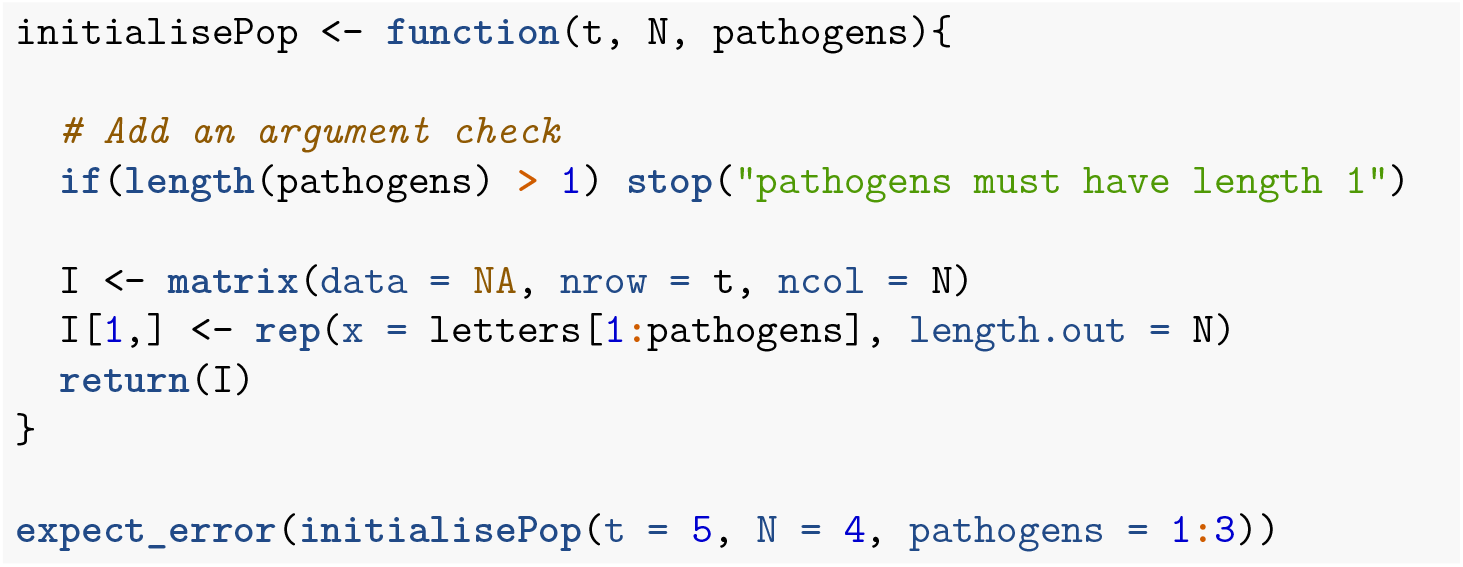

We can similarly check how the code handles a user inputting a vector of numbers to the `t` argument (perhaps thinking it needed a vector of all time points to run). In Code 3, `initialisePop()` does not throw an error if a vector is supplied to `t`. However, `fullSim()` does throw an error if a vector is supplied to `t`. While it is a good thing that `fullSim()` throws an error, the error message is not very informative. If the code that runs before the error is thrown (in this case the `initialisePop()` function) takes a long time, it can also be time consuming to work out what threw the error. It is also a signature of fragile code that the error is coincidental; a small change in the code might stop the error from occurring. To remedy this we can add an additional argument check to `initialisePop()`. Importantly, we then want to check that `fullSim()` errors in the correct place (i.e. in `initialisePop()` rather than afterwards). We can achieve this using the `regexp` argument of `expect_error()` that compares the actual error message to the expected error messages. The test will only pass if the error message contains the string provided.

#### Code 4: Another new definition of the initialisePop() function and a passing test for the fullSimQ function.

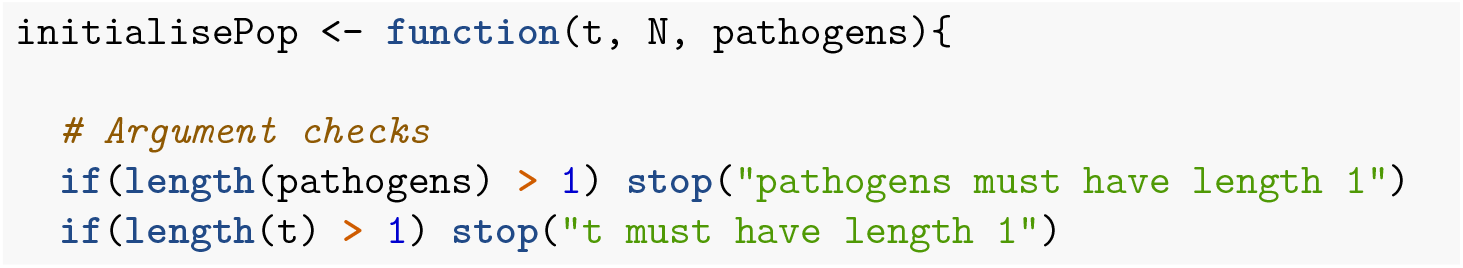
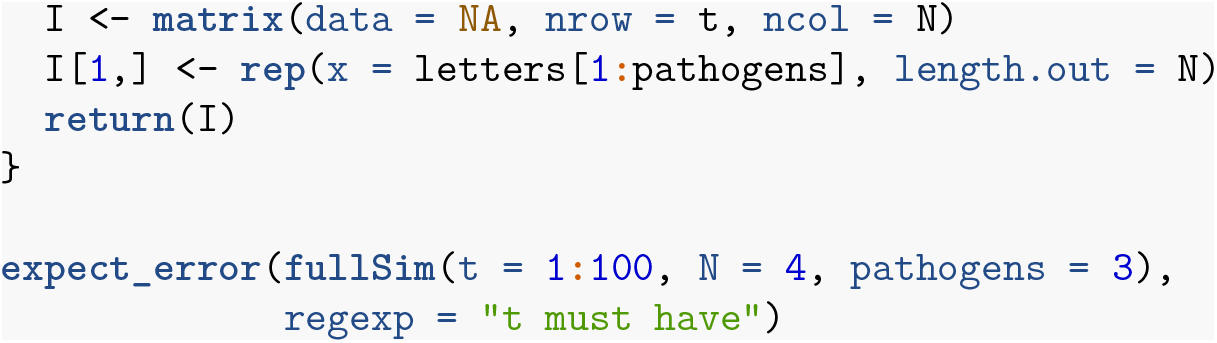

### Test edge cases and special cases

When writing tests it is easy to focus on standard behaviour. However, bugs often occur at *edge cases*—when parameters are at their extrema or at special values. For example, in R, selecting two or more columns from a matrix e.g. `my_matrix[,2:3]` returns a matrix while selecting one column e.g. `my_matrix[,2]` returns a vector. Code that relies on the returned object being a matrix would fail in this edge case.

Similarly, special cases can be triggered with parameter sets that do not match the extrema of parameter space. This is where understanding of the functional form of the model can help. Consider a function `divide(x, y)` that divides `x` by `y`. We could test this function by noting that `y * divide(x, y)` should return `x`. If we write code that tests standard values of `x` and `y` such as `2 * divide(3, 2) == 3` we would believe the function works for nearly all values of division, unless we ever try `y =` 0.

We checked earlier if the `pathogens` argument of `initialisePop()` worked by verifying that the returned population had the correct number of pathogens. However, if we set the `pathogens` argument to be greater than the number of individuals in the population we get a population with `N` pathogens. The function does not therefore pass the test we defined in Code 3.

#### Code 5: initialisePop() does not give a population with the correct number of pathogens if N is less than the number of pathogens.

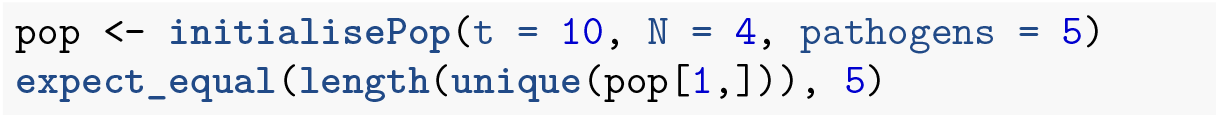

For edge cases like this it may be rather subjective what the correct behaviour should be. It might be appropriate for the function to throw an error or give a warning if the user requests more pathogens than individuals. Here however, we will decide that this behaviour is acceptable. The test above was still useful to highlight this unusual case. As our expected output from the function has changed, we should change our test; we now expect a population with `N` pathogens. We should however retain the test in Code 3 so that we have two tests: one test checks that when `pathogens < N`, the number of unique pathogens in the population is equal to `pathogens`; the other test checks that when `pathogens > N`, the number of unique pathogens is equal to `N`.

#### Code 6: Check that pathogens is equal to N

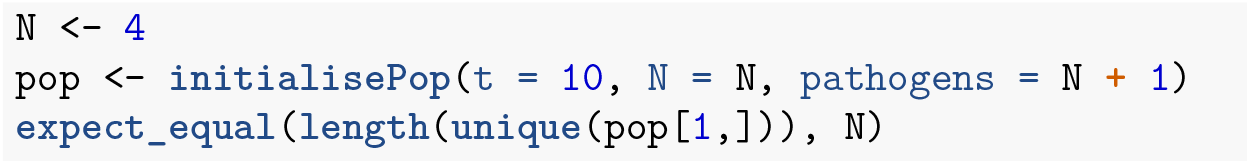

## 7. Unit testing frameworks

Most programming languages have established testing packages. For R, there is the `testthat` package as already discussed. When structuring *R* code as a package, tests should be kept in the directory `tests/testthat`; further requirements to the structure can be found in [22] Chapter 7. All the tests in a package can then be run with `test()` from the `devtools` package [29] or `check()` for additional checks relevant to the package build. If the code is to be part of a package then these tools are essential to run the code within the context of a build environment. These tools also provide a clean environment to highlight if a test was previously relying on objects defined outside of the test script.

Other programming languages have similar testing frameworks. Their specifics differ but the main concept of comparing a function evaluation to the expected output remains the same. In *Julia* there is the `Test` package [30]. The basic structure for tests with this package is demonstrated below. We name the test and write a single expression that evaluates to `TRUE` or `FALSE`. For a *Julia* package, unit tests reside in `test/runtests.jl` and tests are run with `Pkg.test()`.

#### Code 1: Julia test example

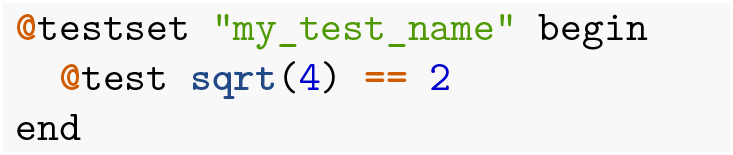

Finally, in *python* we have the `unittest` framework [31]; tests must be written into a class that inherits from the `TestCase` class. The tests must be written as methods with `self` as the first argument. An example test script is shown below. Tests should be kept in a directory called `Lib/test`, and the filename of every file with tests should begin with “`test_`”.

#### Code 2: python test example

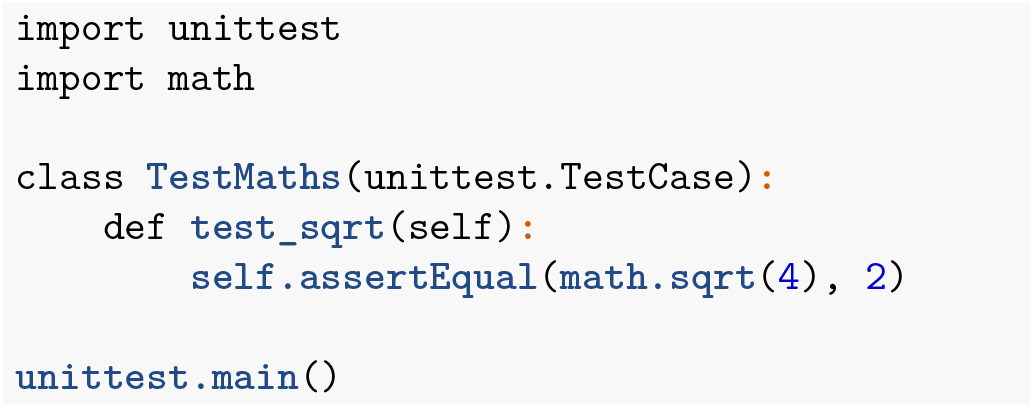

## 8. Continuous integration

If your code is under version control [23,24] and hosted on GitHub, GitLab or BitBucket, you can automate the running of unit tests—also known as *continuous integration*. In this setup, whenever you push code changes from your local computer to the online repository, any tests that you have defined get run automatically. Furthermore these tests can be automated irrespective of changes to your code or tests: since dependencies with other packages and knock-on changes from them into a bug in your code can be automatically found and notified to you by email. There are various continuous integration services such as travis-ci.org, GitHub actions and GitLab pipelines. These services are often free on a limited basis, or free if your code is open source.

We briefly describe the setup of the simplest case usign Travis CI. Setting testing up is very straightforward for *R* code already organised into a package and hosted openly on GitHub. Within your version-controlled folder that contains the *R* code, you add a one-liner file named “ `.travis.yml`” that contains a description of which language the code uses.

### Code 1: A basic travis yml file.

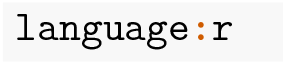

This file can also be created with `use_travis()` from the `usethis` package. You then sign up to travis-ci.org and point it to the correct GitHub repository. To authenticate and trigger the first automatic test you need to make a minor change to your code, commit and push to GitHub. More details can be found in [22] Chapter 14.3.

## 9. Concluding remarks

It is vital that infectious disease models are coded to minimise bugs. Unit testing is a well-defined, principled way we can do this. There are many frameworks that make aspects of unit testing automatic and more informative and these should be used where possible.

The basic principles of unit testing are simple but writing good tests is a skill that takes time, practice and thought. However, ensuring your code is robust and well-tested saves time and effort in the long run and helps prevent spurious results. These qualities are important for the reproducibility of science, but are particularly relevant where code may be shared between individuals or groups, or where you wish to publish your code as a package to be used by others. Our aim in this paper was to demonstrate tailored results for infectious disease modelling. There are number of standard programming approaches to unit testing which would be good followup reading ([22] Chapter 7, [23], [24]). As demonstrated here, it is initially time consuming to program in this way, but over time it will become habitual and both you and the policy-makers who use your models will benefit from it.

## Data Availability

All data available

## 10. Code availability

Please see the fully-reproducible and version-controlled code at http://www.github.com/timcdlucas/unit_test_for_infectious_disease.

## 12. Contributions: CRediT statement

Conceptualization: TCDL TMP

Data curation: TCDL

Formal Analysis: TCDL

Funding acquisition: TDH

Investigation: TCDL

Methodology: TCDL

Project administration: TCDL

Software: TCDL TMP

Validation: TMP

Visualization: TCDL TMP

Writing – original draft: TCDL

Writing – review & editing: TCDL TMP ELD TDH

